# Promoter methylation and gene expression of Pin1 is associated to the risk on Alzheimer’s disease in Southern Chinese

**DOI:** 10.1101/2020.08.23.20172403

**Authors:** Suk Ling Ma, Nelson Leung Sang Tang, Linda Chiu Wa Lam

**Affiliations:** Department of Psychiatry, Faculty of Medicine, The Chinese University of Hong Kong, China; Department of Chemical Pathology, Faculty of Medicine, The Chinese University of Hong Kong, China; Functional Genomics and Biostatistical Computing laboratory, Shenzhen Research Institute, The Chinese University of Hong Kong, China; Laboratory of Genetics of Disease Susceptibility, Li Ka Shing Institute of Health Sciences, The Chinese University of Hong Kong, China

**Keywords:** genetics, Pin1, gene expression, methylation, Alzheimer’s disease, cognitive decline

## Abstract

**Background:** Pin1 is a propyl cis-trans isomerase and it has been associated with age-at-onset of Alzheimer’s disease (AD) and other pathological characteristic of AD. DNA methylation is one of the gene regulation and it might affect the gene expression.

**Objectives:** This study was aimed to examine the correlation between DNA methylation and gene expression of Pin1 and its effect on the risk of AD in a Chinese population.

**Methods:** 80 AD patients and 180 normal controls were recruited in this study and their cognitive function were assessed. Pin1 gene expression and methylation were quantified by real-time RT-PCR and Melting Curve Analysis-Methylation assay (MCA-Meth) respectively.

**Results:** Our finding revealed a positive correlation between methylation and gene expression of Pin1 (p=0.001) and increased Pin1 methylation was predisposed to the risk of AD (p<0.001). CG genotype of Pin1 SNP rs2287839 was associated with higher gene expression of Pin1 (p=0.036) and the effect was only prominent in normal controls as AD patients were already methylated at Pin1 promoter. Furthermore, methylation of Pin1 was associated with better performance in cognition (p=0.018).

**Conclusions:** Our result further supported the involvement of Pin1 in AD and the increased level of Pin1 might be a protective factor for AD.

## 1. INTRODUCTION

Alzheimer’s disease (AD) is the most common neurodegenerative disease in the aging population and the number of affected is expected to increase exponentially due to the increase in lifespan. The pathological characteristics of AD were amyloid plaques and neurofibrillary tangles.

Pin1 is a prolyl isomerase catalyzes the conversion of cis to trans conformation specifically at certain phosphorylated Ser/Thr-Pro motifs [1; 2]. Our previous studies suggested that Pin1 polymorphism rs2287839 was associated with the age-at-onset (AAO) of AD and it modulated the expression of Pin1 by abolishing the binding ability of transcription factor AP4 to Pin1 [3]. However, the study result suggested this polymorphism was not associated with the risk of AD. On the other hand, we and others suggested Pin1 played an important role in pathogenesis of AD by regulating the APP processing and tau dephosphorylation [4; 5].

Epigenetics is defined as the modification of DNA and DNA packaging but do not involve changes to DNA sequence. DNA methylation is one of the major epigenetic mechanism involved in gene regulation and it can directly prevent the binding of transcription factor, resulted in changes in chromatin structure and prevent the access of transcription factor to the promoter region of the gene. The relationship between methylation and gene expression is complex. Inverse relationship between methylation at promoter region and gene expression was reported while a positive association between methylation at gene body and gene expression was suggested [6]. In AD, global hypomethylation was reported in disease-relevant regions of the brain [7]. However, there was also evidence showing global hypermethylation in frontal cortex in AD postmortem cases [8]. In addition, some AD susceptibility genes showed differential methylation pattern [9] between AD patients and normal controls. Hypomethylation of PSEN1 was reported in AD and it was associated with over-expression of PSEN1 [10]. Co-methylation module was observed in brain and blood, suggested blood is a surrogate marker for studying methylation in brain [11]. Our previous study showed UQCRC1 was highly methylated in AD and was positively associated with gene expression of UQCRC1 [12]. Recently, study showed reduction of Pin1 methylation and increased Pin1 expression in Caucasians AD patients [13; 14].

## 2. MATERIALS AND METHOD

### 2.1 Subject recruitment

80 Chinese AD patients (mean age at study = 84.1, SD = 6.1, range 65-96, 83.1% female) with NINCDS-ADRDA diagnosis for probable and possible AD were recruited from the psychogeriatric services of New Territories East Cluster Hospitals in Hong Kong. 180 Chinese older adults (mean age at study = 80, SD = 7.3, range 65-96, 76.5% female) were voluntarily recruited from local elderly social centers residential and hostels for the elderly in Hong Kong. Clinical Dementia Rating (CDR) [15; 16] and Cantonese version of the mini-mental state examination (CMMSE) [17] were performed for all subjects recruited to assess their cognitive function. Subjects with significant sensory deficits or known neurodegenerative and psychiatric disorders were excluded. Informed consent were obtained from the subjects and/or their caregivers. The study has been approved by the Clinical Research Ethics Committee of the Chinese University of Hong Kong.

### 2.2 Genotyping

Genomic DNA was extracted from peripheral blood samples using a DNA extraction kit according to the manufacturer’s instruction (Roche, Nutley, NJ, USA). Pin1 SNP rs2287839 was genotyped as reported earlier [18].

### 2.3 Pin1 gene expression

Peripheral blood samples were stabilized by Trizol (Invitrogen) and RNA was extracted with Trizol according to manufacturer’s instruction. Total RNA was subjected to reverse transcription to cDNA using random hexamer and Superscript III (Invitrogen). The cDNA transcript was amplified by specific primers for Pin1 and quantified using SYBR qPRC kit (KAPA) by LightCycler 480 System (Roche). Melting curves were obtained after amplification cycle 66°C to 95°C. The expression level of Pin1 for each sample was normalized to its level of GAPDH. The relative gene expression levels were calculated by 2-ΔΔCt method [19].

### 2.4 Pin1 methylation

Commercial DNA modification kit was used to perform bisulfite modification for methylation analysis (Zymo Research Corporation). The bisulfated DNA was analysed by Melting Curve Analysis-Methylation assay (MCA-Meth) [20]. Primers for Pin1 were designed using Methprimer software [21] to amplify the promoter region of Pin1 regardless of its methylation status. After bisulfite treatment, CG pairs in the methylated sequence remained unchanged and CG was converted to TG in unmethylated sequence. The melting temperature (the peaks in melting curve) between methylated (M) and unmethylated (U) sequence will have different pattern and it was used to distinguish the samples’ methylation status [20]. The melting curve analysis was performed on LightCycler 480 System (Roche).

### 2.5 Statistical analysis

The distribution of methylation status between AD patients and normal controls were compared using χ^2^ tests. The association between the methylation status and gene expression in the sample set were analysed by non-parametric Spearman’s correlation. Independent t-test was performed to examine the association of methylation status and gene expression level in the group of AD patients and normal controls respectively. Linear and logistic regression analysis were A nominal p-value of < 0.05 was considered as significant association. Statistical analysis were performed using SPSS for windows version 17.0.

## 3. RESULTS

Our result revealed 92% of the AD patients were hypermethylated on Pin1 promoter region and it is significantly higher than that observed in the normal control group (p<0.001) (Table 1). There was no significant association between methylation status of Pin1 and sex, either in whole sample set or stratified by disease affected status.

**Table 1.** Table showing the methylation status in AD patients and normal controls.

|  | **U** | **U/M** | **M** | **p-value** |
| --- | --- | --- | --- | --- |
| Normal controls | 59 (35.1%) | 6 (3.6%) | 103 (61.3%) | <0.001 |
| AD | 1 (1.3%) | 5 (6.7%) | 69 (92.0%) |  |

U: unmethylated; M: methylated.

The gene expression of Pin1 was significantly higher in AD group when compared to normal controls (p < 0.001) (Table 2). We further analysed the correlation between gene expression and methylation in the whole sample set and we identified a positive correlation (Pearson’s correlation coefficient r = 0.213, p=0.001). The gene expression level of Pin1 in subjects with Pin1 promoter methylated was significantly higher than those with unmethylated status (p < 0.001) (Table 2).

**Table 2.** Table showing the correlation between methylation status of Pin1 promoter and gene expression levels in AD and normal controls.

|  | **AD** | **Control** | **p-value** |
| --- | --- | --- | --- |
| % methylation | 92.0% | 61.3% | < 0.001 |
| Relative gene expression<br>(Mean ± SD) | 2.32 ± 2.31 | 0.65 ± 1.57 | < 0.001 |

No significant difference in the genotype distribution of Pin1 SNP rs2287839 between normal controls and AD patients, as reported in our previous study [18]. On the other hand, the gene expression of Pin1 for subjects with CG genotype of rs2287839 was significantly higher than those with GG genotype (p = 0.036) (Table 3). However, the genotype of rs2287839 was not associated with the status of Pin1 methylation. Since both methylation status and genotype of rs2287839 modulated the gene expression of Pin1, we further analysed the correlation between these three variables with linear regression, with gene expression as outcome, genotype of Pin1 SNP and Pin1 methylation status as predictors. The result showed the regression coefficients were 0.139 (methylation status, p = 0.079) and −0.217 (Pin1 SNP, p = 0.006), suggesting both were the modulators for Pin1 gene expression. Interestingly, in control-only analysis, only Pin1 SNP showed significant association with gene expression of Pin1 (p=0.003).

**Table 3.** Table comparing the relative Pin1 gene expression levels between subjects with CG and GG genotypes of rs2287839.

| Genotype of<br>rs2287839 | Mean ± SD | p-value |
| --- | --- | --- |
| CG | 2.13 ± 2.31 | 0.036 |
| GG | 1.08 ± 1.91 | |

The mean score of MMSE for normal subjects was 26.2 and our result showed that methylation of Pin1 was associated with higher score in MMSE score (p=0.018). The effect of methylation on the performance of MMSE was further analysed by logistic regression. Since MMSE score is a continuous variable, the analysis was done by grouping subjects with MMSE score greater or lower than the mean score. The association between methylation and MMSE score remained significant after controlled for age and sex (p=0.023, 95% CI = 0.479 - 0.946). However, such association between cognitive decline and methylation was not observed in AD group.

## 4. DISCUSSION

Genome-wide study on gene expression and methylation status suggested the correlation between methylation and gene expression could be positive or negative and the correlation is predominantly negative [22]. In our current study, positive correlation between methylation status and gene expression of Pin was observed. Furthermore, the increased level of methylation and gene expression of Pin1 was associated with the risk of AD and cognitive decline in normal controls.

In AD patients, reduced activity and gene expression of Pin1 was reported and studies showed Pin1 is highly oxidized in hippocampus from AD [23]. On the other hand, there was study showed the reverse, reporting increased gene expression and activity in AD patients when compared to normal controls [24]. The discrepancies on the level of Pin1 reported in AD patients’ brain might be due to the difference of the stage of disease. Tau phosphorylation is one of the pathologic hallmarks for AD and previous study showed Pin1 binds to phospho-Tau and stimulates tau dephosphorylation at Cdk5-mediated phosphorylation sites [4]. Wang et al proposed that in the presence of increased level of phosphorylated tau in AD patients, Pin1 activity was increased due to a compensatory up-regulation mechanism and this resulted in the increase in gene expression of Pin1 [24]. On the other hand, increased neuronal loss in patients’ brain is observed as the disease progressed to a later stage and this might result in reduced Pin1 activity as the number of cells expressing Pin1 is decreased when compared to the early stage of AD [23].

In our previous study, we found CG genotype of rs2287839 was associated with the abolishment of the binding activity of AP4 to Pin1 and therefore AP4 can no longer suppress the activity of Pin1 promoter expression [18]. In the current study, we identified CG genotype was associated with higher gene expression of Pin1, which is in line with our previous study. CG genotype of rs2287839 was associated with a 3-year delay in average AAO and the increased Pin1 expression might account for the protective effect in delaying the onset of the disease. On the other hand, genotypes of rs2287839 were not associated with the susceptibility of AD and no significant increase in gene expression of Pin1 was observed in AD patients with CG genotype. Since 92% of the AD patients were highly methylated at Pin1 and methylation was associated with increased in gene expression of Pin1, the influence of genotypes of rs2287839 on Pin1 gene expression was expected not significant when compared to the effect of the highly methylated status of Pin1. DNA methylation not only affects disease susceptibility, previous study reported the negative correlation between LINE1 methylation and MMSE score [25]. In our study, we observed positive correlation between Pin1 methylation and MMSE score. When Pin1 was methylated, the protective effect of the increased gene expression of Pin1 might account for the better performance in cognition.

There was discrepancy between the result from our current study and Arosio et al [13]. We identified positive correlation between DNA methylation status and gene expression level of Pin1 while a negative correlation was reported by Arosio et al. Pin1 level might increase when the disease became more severe, due to the compensatory mechanism and the reducing number of neuronal cells would reduce the gene expression of Pin1. In addition, there is causal relationship between aging and DNA methylation [26] and it is unclear how is the correlation between the changes in DNA methylation due to aging and the pathology of AD. The contrasting result between our current findings and Arosio et al might be due to the different timing and staging of AD when the patients’ samples were collected and analysed. Further longitudinal study at different time point during the aging process will be required to better elucidate the relationship between methylation and gene expression of Pin1 on the pathogenesis of AD.

## CONCLUSION

Our study showed there is positive correlation between Pin1 methylation and gene expression. The increased level of methylation and gene expression of Pin1 was associated with the risk of AD.

## Data Availability

The data is available on request.

## ETHICS APPROVAL AND CONSENT TO PARTICIPATE

The study has been approved by the Clinical Research Ethics Committee of the Chinese University of Hong Kong.

## HUMAN AND ANIMAL RIGHTS

No animals were used in this research. All humans research procedures were accordance with the Declaration of Helsinki.

## CONFLICT OF INTEREST

The authors declare no conflict of interest, financial or otherwise.

## ACKNOWLEDGEMENTS

The study was supported by the Direct Grant from the Chinese University of Hong Kong.

## Notes

### Competing Interest Statement

The authors have declared no competing interest.

